# Early Detection of SARS-CoV-2 Omicron BA.4/5 in German wastewater

**DOI:** 10.1101/2022.07.27.22278003

**Authors:** Alexander Wilhelm, Jens Schoth, Christina Meinert-Berning, Daniel Bastian, Sandra Ciesek, Burkhard Teichgräber, Thomas Wintgens, Frank-Andreas Weber, Marek Widera

## Abstract

Wastewater-based SARS-CoV-2 epidemiology (WBE) has been established as an important tool to support individual testing strategies. Omicron sub-variants BA.4/5 have spread globally displacing the predeceasing variants. Due to the severe transmissibility and immune escape potential of BA.4/5, early monitoring was required to asses and implement countermeasures in time.

In this study, we monitored the prevalence of SARS-CoV-2 BA.4/5 at six municipal wastewater treatment plants (WWTPs) in the Federal State of North-Rhine-Westphalia (NRW, Germany) in May and June 2022. Initially, L452R-specific primers/probes originally designed for SARS-CoV-2 Delta detection were validated using inactivated authentic viruses and evaluated for their suitability to detect BA.4/5. Subsequently, the assay was used for RT-qPCR analysis of RNA purified from wastewater obtained twice a week at six WWTPs. The occurrence of L452R carrying RNA was detected in early May 2022 and the presence of BA.4/5 was confirmed by variant-specific single nucleotide polymorphism PCR (SNP-PCR) targeting E484A/F486V. Finally, the mutant fractions were quantitatively monitored by digital PCR confirming BA.4/5 as the majority variant by 5th June 2022.

In conclusions, the successive workflow using RT-qPCR, variant-specific SNP-PCR, and RT-dPCR demonstrates the strength of WBE as a versatile tool to rapidly monitor variant spreading independent of individual test capacities.

## 1. Introduction

The severe acute respiratory syndrome coronavirus type 2 (SARS-CoV-2) is the causative agent of the coronavirus disease 2019 (COVID-19). SARS-CoV-2 was identified in December 2019 in the Chinese metropolis of Wuhan and rapidly evolved into a global pandemic [1,2]. During the evolutionary process, nucleotide polymorphisms might arise in the SARS-CoV-2 genome potentially affecting the viral transmissibility, susceptibility to monoclonal antibodies (mAb) and antibodies from convalescent as well as vaccine-elicited sera [3]. In recent months, several lineages of the SARS-CoV-2 Omicron variant (B.1.1.529) including BA.1 and BA.2 have prevailed over the preceding Delta variant [4]. Compared to the predecessor variants, BA.1 and BA.2 were shown to exert severe immune escape to naturally acquired or vaccine-elicited neutralising antibodies [5-9]. The two most recent variants BA.4 and BA.5 have high similarity to BA.2 but carry exclusive mutations including Δ69/70; L452R, F486V, and Q493 in the spike protein, which is important for cellular entry but also immune evasion [10,11]. In particular, substitutions in Spike L452R/Q/M are associated with a high reproduction number. BA.4 and BA.5 were shown to severely escape antibodies elicited by early Omicron infection indicating advanced resistance to previous immunity [12,13]. Furthermore, early published data reports more efficient spread in lung cells and potential higher pathogenicity of BA.4/5 when compared with BA.2 [11]. Hence, due to the severe immune escape, high transmissibility, and unknown pathogenic potential, early monitoring of BA.4/5 is required to consider the regulation of local countermeasures.

Using molecular reverse transcriptase quantitative polymerase chain reaction (RT-qPCR) analysis the detection of SARS-CoV-2 gene fragments in wastewater was shown to correlate with the COVID-19 prevalence in the catchment area of the sewage treatment plant [14,15]. A wide variety of advanced detection methods including RT-qPCR, digital (droplet) PCR (d(d)PCR), and genomic sequencing (NGS) were described to detect and quantify variant specific RNA fragments [16-22]. For all approaches, substitutions absent in the predecessing variant were shown as suitable target for molecular detection.

In this study, we demonstrate a step-wise coordinated workflow using the advantages of multiple PCR detection methods for rapid tracking of the SARS-CoV-2 Omicron BA.4/5 variant. Using pre-existing L452R-PCR and a variant-specific single nucleotide polymorphism PCR targeting E484A/F486V, we describe a rapid and cost effective monitoring approach, might be applicable to monitor future pandemic waves.

## 2. Materials and Methods

### 2.1 Sewage sampling

Wastewater samples were collected at six municipal waste water treatment plants (WWTPs): Klaeranlage Emschermuendung (KLEM), Dortmund-Scharnhorst (DoS), Dortmund-Deusen (DoD), Bottrop (BOT), Dinslaken (DIN), and Duisburg-Alte-Emscher (DAE), all located in North-Rhine Westphalia (NRW) / Germany (Figure 1). Key properties of the WWTPs operated by the public German water board Emschergenossenschaft and Lippeverband are summarized in Table 1. Flow-proportional 24-h composite samples were collected after the grit chamber at the WWTP inlet using an installed autosampler.

**Table 1.** Key properties of the six wastewater treatment plants sampled in this study. Data was obtained from ELWAS-WEB, an electronic water management system for administration in NRW (accessed 29.06.2022).

| WWTP | Acronym | Nominal number of connected residents | Population equivalent | Annual wastewater flow in 2020 [m<sup>3</sup>/a] |
| --- | --- | --- | --- | --- |
| Emschermuendung | KLEM | 906,222 | 426,173 | 348,703,426 |
| Dortmund-Scharnhorst | DoS | 113,439 | 45,342 | 12,192,038 |
| Dortmund-Deusen | DoD | 399,425 | 185,144 | 47,716,171 |
| Bottrop | BOT | 732,816 | 678,818 | 131,203,662 |
| Duisburg Alte Emscher | DAE | 242,172 | 133,535 | 35,807,933 |
| Dinslaken | DIN | 56,812 | 19,157 | 3,812,870 |

**Figure 1.**
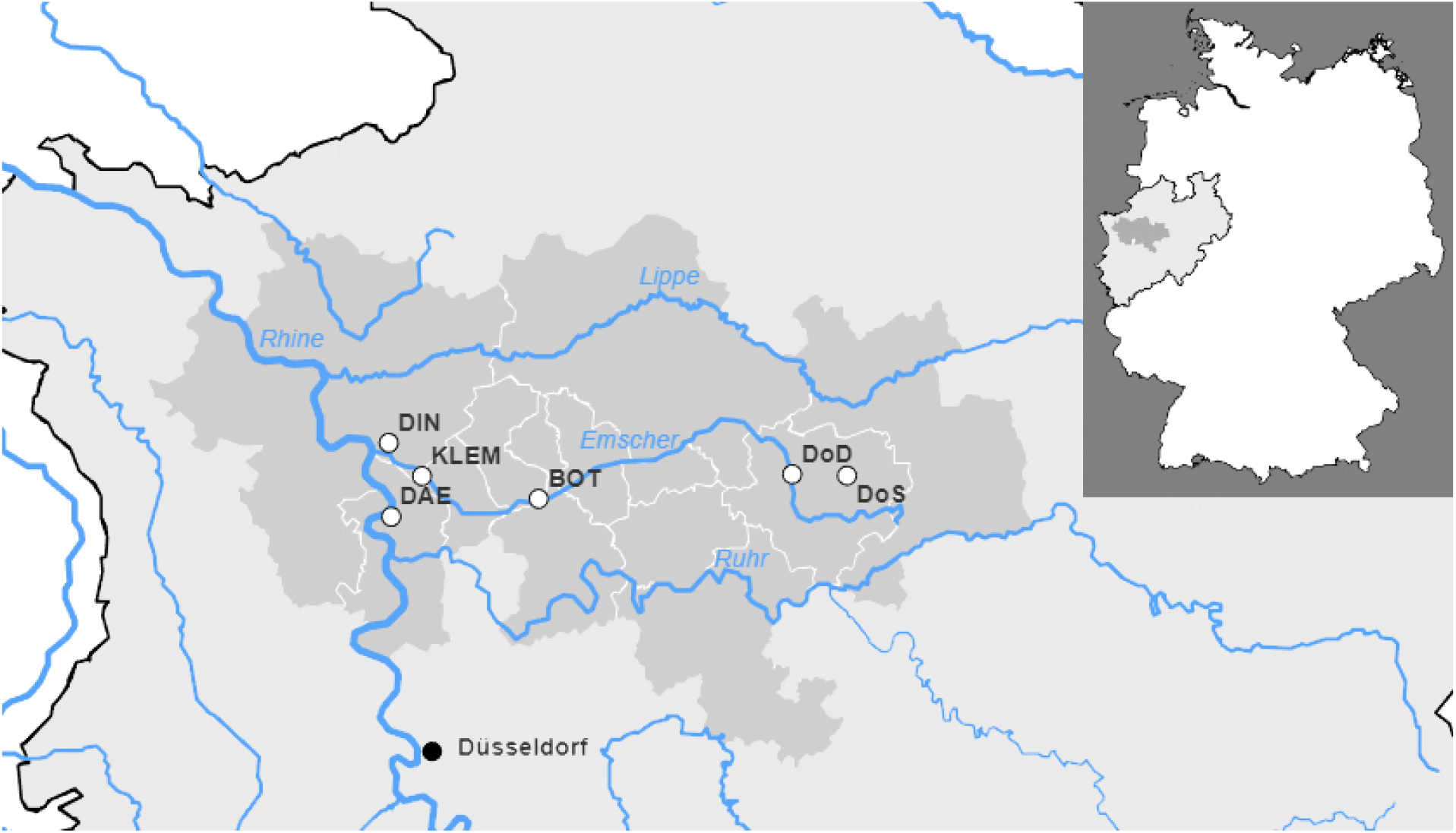
The wastewater treatment plants monitored as part of this study. Regional representation of the municipal districts (dark gray) connected to the respective wastewater treatment plants in the German state of North Rhine-Westphalia. The sewage treatment plants are shown as white circles with black borders. Rivers are shown in blue, federal state borders in black. Cities are shown as black circles for further orientation. A general map of Germany to illustrate the overall view, indicating the colors, is given at the top right.

### 2.2 Sample processing and RT-qPCR quantification of viral RNA

Sample processing and RT-qPCR quantification of viral RNA was performed as described previously [16]. Accordingly, 100 ml of each sample were filtered through electronegative membrane filters (0.45 µm pore size, Merck Millipore, Darmstadt, Germany) at a pressure of 6 bar using Nitrogen gas. After filtration, the filter was cut and placed onto Lyses Tubes J (Analytik Jena, Jena, Germany). After addition of 1 ml DNA/RNA shield reagent (Zymo Research Europe, Freiburg im Breisgau, Germany) the samples were processed in a speed mill (SpeedMill plus, Analytik Jena) at 50 Hz for 2 min and centrifuged subsequently. Total RNA of the samples was isolated using the automated purification system InnuPure C16 touch (Analytik Jena, Jena) and the innuPREP AniPath DNA/RNA Kit IPC16 according to the manufacturer’s instructions. Extracted RNA was used directly for RT-qPCR and subsequently stored at -80°C.

The initial screening for the presence of the Omicron variant was performed at the cooperative laboratory of Emschergenossenschaft and Lippeverband in Essen, which is operated together with Ruhrverband, by analysis of the following mutation: L452R using the QuantiNova Pathogen +IC Kit (Qiagen, Hilden, Germany) on a qTOWER^3^ real-time-thermocycler (Analytic Jena, Jena, Germany) according the manufacturers’ instructions. The L452R assay specificity and sensitivity for the detection of L452R-carrying SARS-CoV-2 variants were evaluated using inactivated authentic viruses and patient derived swab samples (Supplementary Figure 1).

### 2.3 Sample processing and RT-qPCR quantification of viral RNA

Wastewater samples were processed using an optimized workflow based on the previously described 4S method [23]. 40 mL of wastewater samples were poured into a 50 mL tube containing 9.35 g sodium chloride and 400 µL of TE buffer (1 M Tris, 100 mM EDTA, pH 7.2).

Samples were agitated until all sodium chloride was dissolved and heat-inactivated at 70°C for 45 min. Each sample was filtered twice through a 5 µM filter and mixed with 70% ethanol in a 1:1 ratio. Subsequently, RNA was isolated using an adapted protocol of the Wizard Enviro TNA Kit (Promega, Walldorf, Germany). RNA was eluted 50 µL RNA-free water for subsequent analysis using the QIAcuity OneStep Advanced Probe Kit (Qiagen, Hilden, Germany) performed on a QIAcuity Digital PCR System (Qiagen, Hilden, Germany). Primer and probe sequences are described in Supplementary Table 1.

### 2.4 SARS-CoV-2 variant-specific single nucleotide polymorphism PCR (SNP-PCR)

After initial screening for the presence of L452R, a variant-specific single nucleotide polymorphism PCR targeting E484A/F486V was performed to confirm BA.4/5. The proprietary primer and probes (SARS-CoV-2 VirSNiP Mutations Assay, Cat.-No. 53-0839-96) were purchased by TIB Molbiol Syntheselabor GmbH (Berlin, Germany). After cycling, a melting curve analysis was performed using a heating rate of 0.2°C / second. A specific melting peak predecessor variants, indicates the specific SARS-CoV-2 variant of concern.

### 2.5 Specificity testing using authentic SARS-CoV-2 isolates

Nasopharyngeal swabs derived authentic SARS-CoV-2 isolates and original sample material were used for specificity testing as described previously [9]. All cell culture work involving infectious SARS-CoV-2 was performed under biosafety level 3 (BSL-3) conditions. Sample inactivation for further processing was performed with previously evaluated methods [24].

### 2.6 Epidemiological data

Epidemiological data on SARS-CoV-2 cases including the variant specific portions in NRW were obtained from the official data repository of the Federal Robert Koch Institute (RKI) in charge of public health surveillance.

## 3 Results

### 3.1. Wastewater surveillance allows early detection of SARS-CoV-2 Omicron variants BA.4/5

Wastewater samples were collected at six municipal waste water treatment plants: Klaeranlage Emschermuendung (KLEM), Dortmund-Scharnhorst (DoS), Dortmund-Deusen (DoD), Bottrop (BOT), Dinslaken (DIN), and Duisburg-Alte-Emscher (DAE), all located in North-Rhine Westphalia (NRW) / Germany (Figure 1).

To monitor the occurrence of BA.4/5, we used a primer/probe set that was already available for the detection of SARS-CoV-2 Delta (Supplementary Table 1). The assay was validated using several inactivated SARS-CoV-2 variants of concern (VoCs) and patient derived swab samples for subsequent RT-qPCR analysis in a decentral laboratory near the WWTPs.

Wastewater samples were initially monitored for total viral load (N1/N2) by RT-qPCR showing an increasing viral load in all observed WWTP beginning on 1st June 2022 (Figure 2A). Concomitantly to this increase, lower ct-values for the BA.4/5 characteristic substitution L452R were observed using a target specific PCR assay yielding FAM signal for L452R and Cy5 signal for L452. Starting with a peak on 23rd of May in wastewater treatment plant Dinslaken (DIN), and on 25th of May for wastewater treatment plant Dortmund-Deusen (DoD), Dortmund-Scharnhorst (DoS), Emschermuendung (KLEM), Duisburg-Alte-Emscher (DAE) and on the 1^st^ of June in Bottrop (BOT) (Figure 2B). A preceding increase of the ct-values for L452 could already be observed on 18/05/2022 at most wastewater treatment plants. At DoS, this increase was observed with a delay of 5 days on 23/05/2022 (Figure 2B). A first rough estimation of the relative proportions of the L452R mutant based on the reciprocal ct-values for L452/L452R revealed a more than 50% L452R fraction on 25/05/2022 for DoD and DAE, on 30/05/2022 for DoS, and on 01/06/2022 for KLEM, DIN, DAE and BOT (Figure 2C). However, for the treatment plants DoD and DAE, a strong drop was observed in the follow-up measurement, which may be due to the non-absolute copy number determination of the RT-qPCR method.

**Figure 2.**
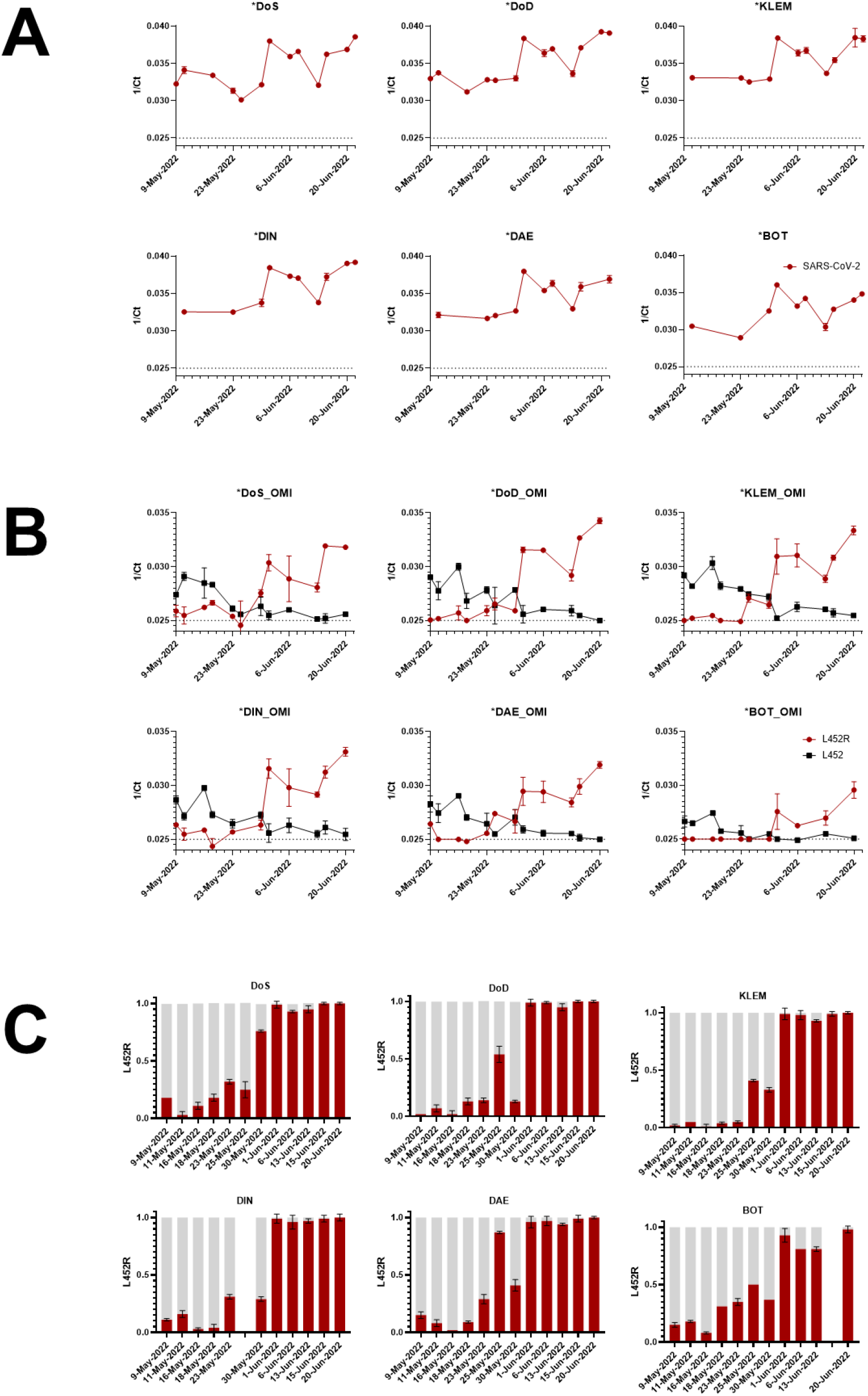
Monitoring of SARS-CoV-2 viral RNA fragments in six different wastewater treatment plants. Wastewater samples were analysed by RT-qPCR for A) the presence of SARS-CoV-2 viral load using N1/N2 targets, B) Omicron BA.4/5 characteristic mutation L452R in SARS-CoV-2 Spike. The corresponding reciprocal ct-values are illustrated for each WWTP. C) Red (L452R) and grey (L452) bars represent the ratio between BA.4/5 and non-BA.4/5 fractions estimated by calculating the ct-values for both targets. For technical reasons, no data are available for DIN from 25.05.2022 and for BOT from 15.06.2022.

This first evidence of the non-exclusive BA.4/5 substitutions (L452R) was further subjected to a variant-specific single nucleotide polymorphism PCR targeting E484A/F486V.

### 3.2. Confirmation of Omicron BA.4/5 by variant-specific single nucleotide polymorphism PCR

To confirm that the L452R-based detection definitely originated from SARS-CoV-2 BA.4/5, a variant-specific single nucleotide polymorphism PCR, originally developed for individual testing, was performed. A preceding validation of the assay approved its use also for the detection of substitution in wastewater samples (Supplementary Figure 1). Subsequently, consecutive samples collected at several sites were measured with this assay. The melting curve analysis of the amplicon products clearly confirmed the presence of a variant carrying E484A/F486V, which is exclusively found in BA.4/BA.5 (Figure 3). In agreement with qRT-PCR derived data, the comparison of both peaks E484A (present in BA.1/2) and E484A/F486V (specific for BA.4/5) [25] revealed a shift towards E484A/F486V confirming the presence of BA.4/5. A concomitant decrease of the E484A-specific peak confirmed this assumption.

**Figure 3.**
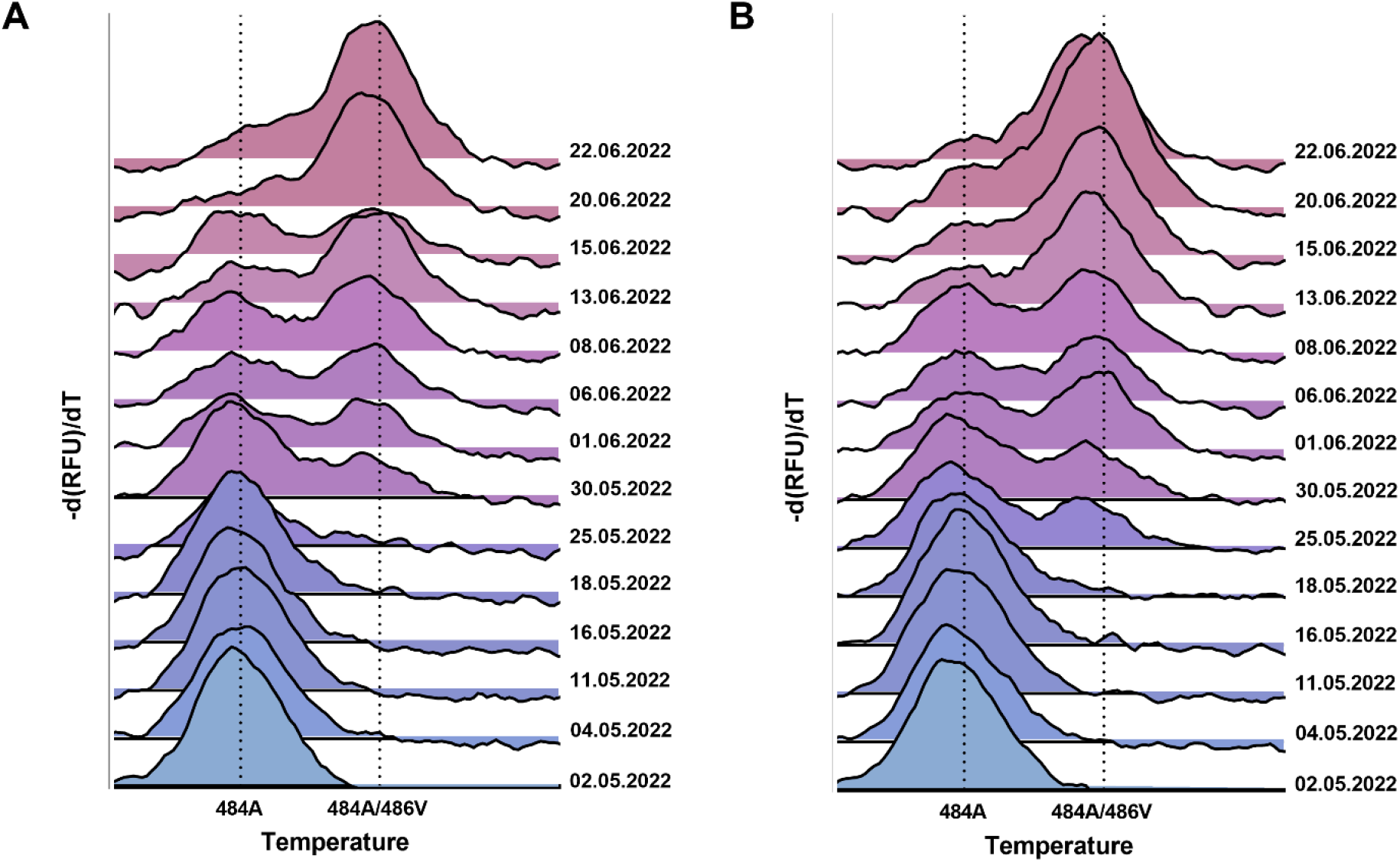
Confirmation of E484A/F486V substitution by Melting Curve PCR analysis. A variant-specific single nucleotide polymorphism PCR (SNP-PCR) targeting E484A/F486V for performed with RNA derived from the two WWTPs A) KLEM and B) DoD. The peak on the left (E484A) shows the specific melting temperature of an amplicon containing E484A representing non-BA.4/BA5 SARS-CoV-2 RNA. Due to the low binding caused by mismatch, a lower temperature is required compared to E484A/F486V with more efficient probe match. The color scheme is for orientation only and does not correlate with the proportion of the mutant.

### 3.3 Tracking the mutation fraction using digital PCR

Since the assay for L452R was already predesigned to detect the SARS-CoV-2 Delta variant and evaluated for high specificity and sensitivity for BA.4/BA.5 (Supplementary Figure 1), this target was used as an applicable surrogate marker to track the course of mutant fractions over time. As shown in Figure 4A, only a minor difference comparing the national and NRW specific 7-days incidence was observed during the study period (Figure 4A, left panel). Following individual testing, Omicron BA4./5 has become the dominant variant in Germany on a national scale on 12th July 2022 (Figure 4A, right panel).

**Figure 4.**
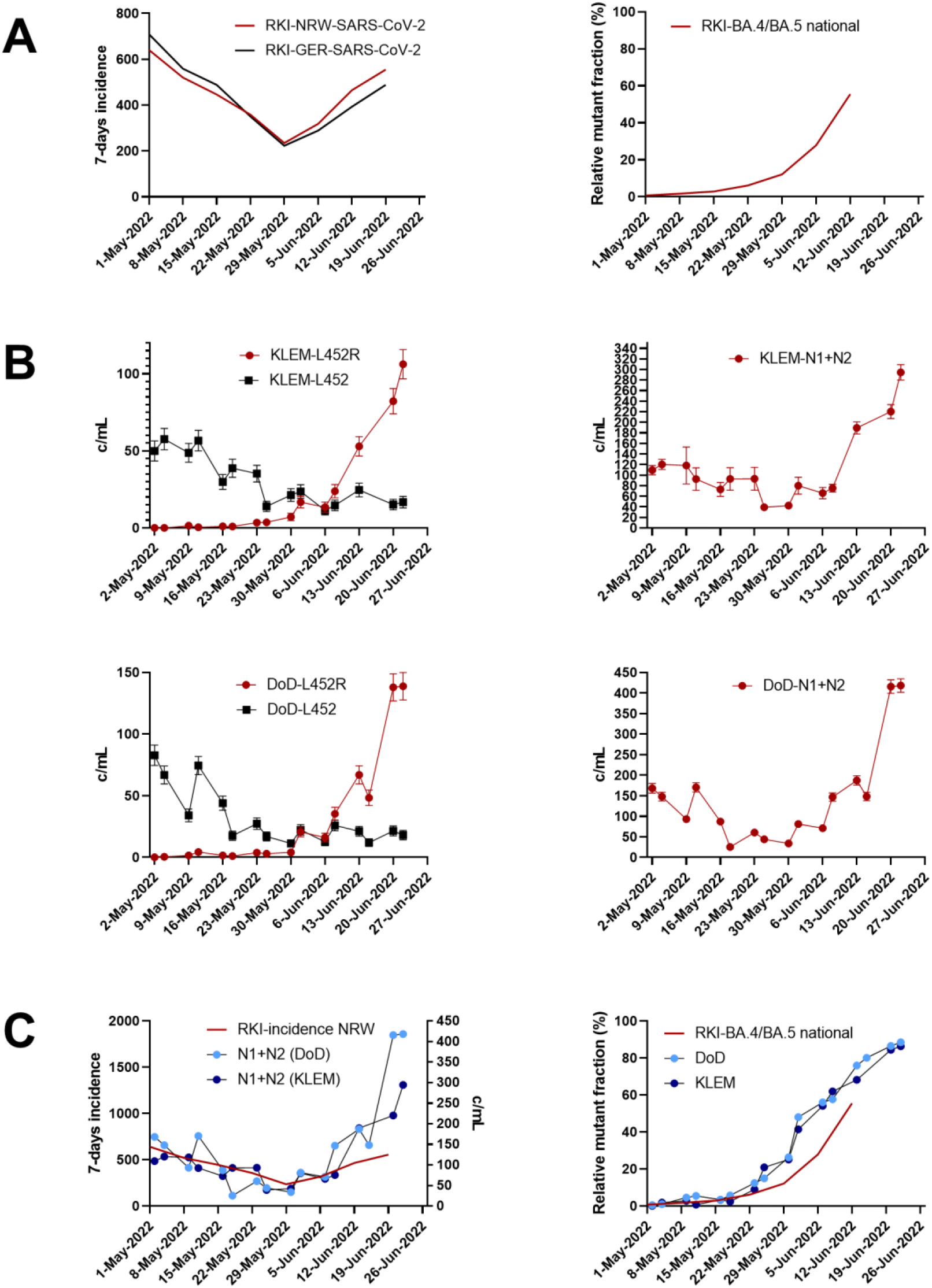
Tracking of SARS-CoV-2 BA.4/BA.5 specific mutant fraction of L452R using digital PCR. A) 7-days incidence (left panel) and the SARS-CoV-2 Omicron BA.4/5 mutant fraction (right panel) for Germany are indicated. Epidemiological data are based on individual testing and were obtained from official data repository of the German Federal Robert Koch Institute (RKI). B) SARS-CoV-2 Omicron BA.4/5 and non-BA.4/5 detection targeting the substitution L452R (left panel) for WWTPs DoD and KLEM. Total SARS-CoV-2 levels in WWTPs DoD and KLEM were quantified using a SARS-CoV-2 specific N1/N2 assay (right panel) C) Overlay of the 7-days incidence in NRW compared to detected SARS-CoV-2 levels in wastewater of WWTPs DoD and KLEM (left panel). Overlay of the relative mutant fraction of SARS-CoV-2 Omicron BA.4/5 as provided by RKI compared to the relative fraction of L452R determined in wastewater using RT-dPCR (right).

The dPCR-derived findings suggest that the Omicron variants BA.4/5 occurred in the WWTP catchment area at rapidly rising fraction (>40%) on the 25th of May in both WWTPs KLEM and DoD (Figure 4B). The decrease of L452 specific signal clearly indicated the displacement of SARS-CoV-2 BA.2.

Ahead of available public-health data at this time point, the relative increase of the mutant fraction over time could be quantitatively monitored confirming Omicron as the predominant variant on 26th June 2022. In agreement with public health data, the dPCR-based quantification of total SARS-CoV-2 RNAs (N1/N2) was precisely resembling the 7-days incidence. Of note, the strong increase in total SARS-CoV-2 RNA measured with a N1 and N2 specific assay strongly correlated with the dominance of L452R. Our data on the current BA.4/5 monitoring demonstrate the strength of decentralized wastewater monitoring.

## 4. Discussion

Data from wastewater-based SARS-CoV-2 epidemiology (WBE) was shown to correlate with the COVID-19 prevalence in the catchment area of the respective sewage treatment plants [14,15]. Depending on the WWTP, catchment area, and the specific ratio of commuters or permanent residents, WBE may even provide time advance in reporting of several days compared to clinical findings. Hence, WBE can effectively support individual testing strategies and becomes even more important in times of declining testing willingness. In addition, WBE might be used to monitor the emergence of novel variants of concern (VoC) already at early stages. Previous studies have shown that measuring the concentration of variant specific SARS-CoV-2 RNA in wastewater can efficiently track regional variant dynamics [16].

Recently, the SARS-CoV-2 variant Delta was predominant until the end of 2021 but was rapidly displaced. In a study performed by the COVIDready consortium in December 2021, we were able to identify the occurrence of the novel SARS-CoV-2 variant Omicron (BA.1) wastewater using a fast and efficient decentralised workflow [16]. The hallmark of this workflow was that previously existing detection assays, such as those for the detection of the K417N substitution originally validated for the detection of the Beta variant, were immediately available for the detection of new variants harboring the identical substitution. Importantly, K417N was present in the Omicron BA.1 and BA.2 subvariants, but not in the previously dominant Delta variant. Hence, by simultaneous detection of the parental variant, a continuous transition of both variants could be tracked over time and quantified by digital PCR, not only in a relative but also in an absolute manner.

An initial screening using already available PCR assays allows early monitoring, however, careful evaluation of the assays for specificity is highly recommended. Many commercially PCR assays are able to detect the newly emerging variant, but may give false positive signals for other variants. For individual testing, these weak false positive signals are negligible, as the sample usually contains a single variant. The wastewater matrix, however, is much more complex, as the sum of all variants within the catchment area of a WWTP is covered simultaneously. The WWTP KLEM, for example, is connected to more than 900,000 nominal residents (Table 1).

The current detection of BA.4/BA.5 using a PCR assay detecting L452R originally developed for Delta detection, but not targeting Omicron variants BA.1 and BA.2, again proved the effectiveness of the decentral workflow. Initially, primers and probes originally designed for the detection of the SARS-CoV-2 Delta-specific L452R-substitution were rapidly validated using inactivated authentic viruses and evaluated for their suitability to detect BA.4/5 (Supplementary Figure 1). In addition to the detection of characteristic but not exclusive mutations, the newly emerging variant needs to be confirmed and identified which was so far performed by cost ad time intensive sequencing methods. In this study, the initial workflow was further developed and the verification of the emerging SARS-CoV-2 variant was replaced by a rapidly available, inexpensive and excellent performing single nucleotide polymorphism PCR (SNP-PCR). Compared to the preceding variant, only a few substitutions in the spike gene including L452R and F486V might be used for discrimination. Using a variant-specific SNP-PCR targeting E484A/F486V, we confirmed the presence of SARS-CoV-2 Omicron BA.4/5. Because of their rapid availability, variant specific melting curve-based PCRs have proven to be a suitable method for rapid confirmation. Of note, the time course was in excellent agreement with the dynamics quantified via dPCR on the basis of the L452R ratios. However, in order to finally distinguish BA.4 from BA.5, NGS-based sequencing would still have to be carried out. However, SNP-PCR allows rapid confirmation of previously available assays without the time delay that is inevitable due to the processing time of a sequencing.

We conclude that a coordinated PCR-based workflow might serve as a robust and sensitive early warning system in pandemic control. We recommend to implement such a coordinated workflow in German wastewater-based epidemiology as a complementary measure in addition to individual testing strategies. Using this established workflow synergistically with NGS, we will be able to rapidly adapt to and track the spread and dynamics of new SARS-CoV-2 variants and future pandemics.

## Data Availability

All data produced in the present study are available upon reasonable request to the authors.

### Appendix

**Supplementary Figure 1.**
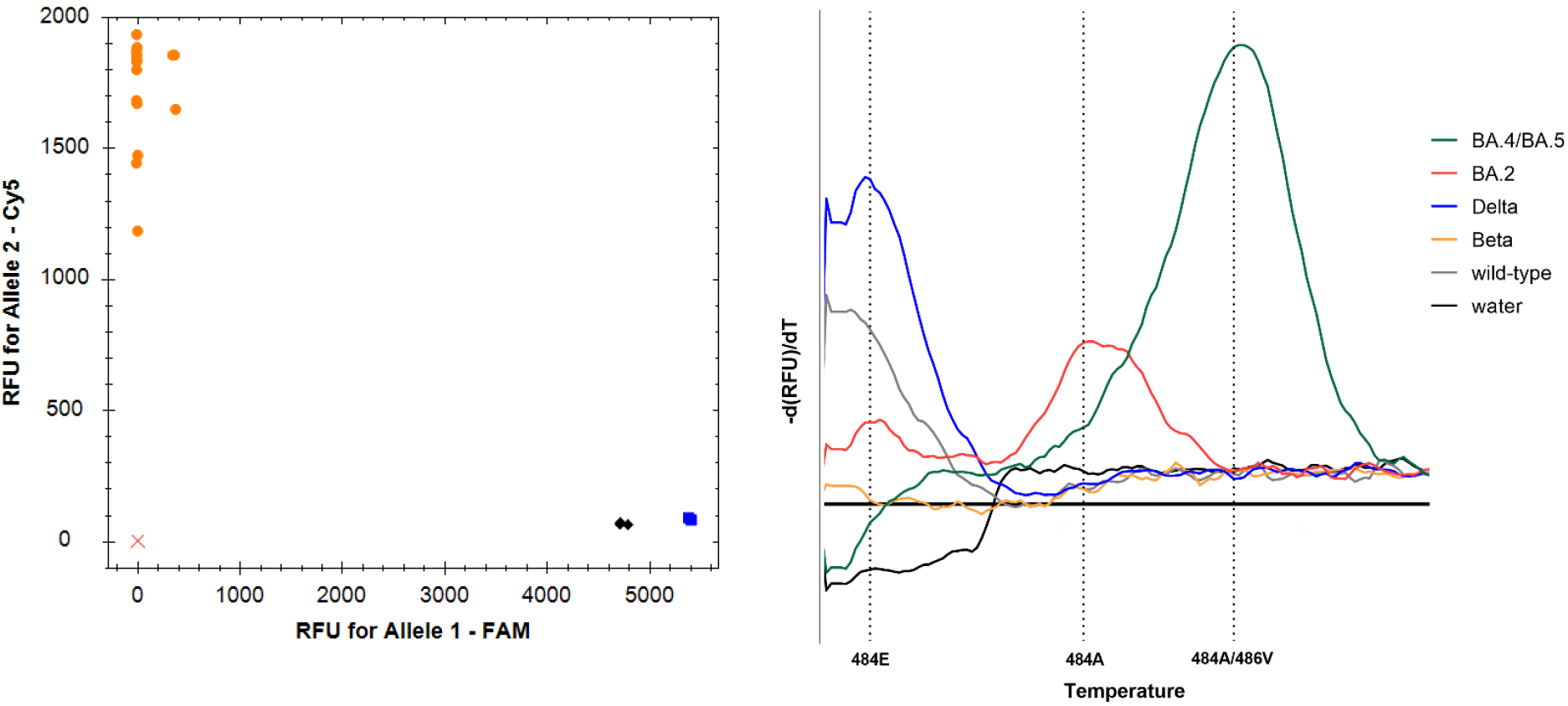
Specificity and sensitivity of PCR assays used in this study. A) Specificity and sensitivity of L452R Assay. Samples spiked with inactivated SARS-CoV-2 variants and patient derived swab samples were used for assay validation (n=3). SARS-CoV-2 VoCs Delta and BA.4/BA.5 were exclusively detected by the FAM-labelled probe detecting L452R (Delta shown in black and BA.4/BA.5 depicted in blue). Parental SARS-CoV-2, Alpha, and Beta were exclusively detected by the Cy5-labelled probe detecting L452 (all L452 carrying variants are depicted in orange). The red cross represents the water-control. B) Specificity and sensitivity of variant-specific single nucleotide polymorphism PCR. Samples spiked with inactivated SARS-CoV-2 variants and patient derived swab samples were used for assay validation.

## Author Contributions

Alexander Wilhelm: Investigation, Visualization, Methodology, Writing-Reviewing and Editing. Jens Schoth: Writing-Reviewing and Editing, Visualization, Resources. Christina Meinert-Berning: Investigation, Writing-Reviewing and Editing. Daniel Bastian: Visualization, Writing-Reviewing and Editing. Sandra Ciesek: Writing-Reviewing and Editing, Supervision, Project administration. Burkhard Teichgräber: Project administration, Supervision, Resources, Writing-Reviewing and Editing. Thomas Wintgens: Writing-Reviewing and Editing, Supervision, Project administration. Frank-Andreas Weber: Conceptualization, Writing-Reviewing and Editing, Funding acquisition, Project administration. Marek Widera: Investigation, Visualization, Writing-Original draft preparation, Funding acquisition, Conceptualization, Supervision, Validation.

## Acknowledgments

We are thankful for the numerous donations to the Goethe-Corona-Fund of the Goethe University Frankfurt (AW, MW) and the support of our SARS-CoV-2 research. Furthermore, we would like to thank Christiane Pallas, Irina Jakobsche, Julia Banholzer, and Joanna Landgraf for their support conducting the analysis of the wastewater samples. We thank the employees of the water boards Emschergenossenschaft and Lippeverband (EGLV), and Ruhrverband, for their participation in the sampling campaign and analyses.

